# The Effect of a Post-Bronchodilator FEV_1_/FVC *<* 0.7 on COPD Diagnosis and Treatment: A Regression Discontinuity Design

**DOI:** 10.1101/2024.08.05.24311519

**Authors:** Alexander T. Moffett, Scott D. Halpern, Gary E. Weissman

**Affiliations:** Division of Pulmonary, Allergy, and Critical Care Medicine, Department of Medicine, University of Pennsylvania, Philadelphia, PA, USA; Palliative and Advanced Illness Research (PAIR) Center, University of Pennsylvania, Philadelphia, PA, USA; Leonard Davis Institute of Health Economics, University of Pennsylvania, Philadelphia, PA, USA; Department of Biostatistics, Epidemiology and Informatics, University of Pennsylvania, Philadelphia, PA, USA; Department of Medical Ethics and Health Policy, University of Pennsylvania, Philadelphia, PA, USA

## Abstract

**Background:** Global Initiative for Chronic Obstructive Lung Disease (GOLD) guidelines recommend the diagnosis of chronic obstructive pulmonary disease (COPD) only in patients with a post-bronchodilator forced expiratory volume in 1 second to forced vital capacity ratio (FEV_1_/FVC) less than 0.7. However the impact of this recommendation on clinical practice is unknown.

**Research Question:** What is the effect of a documented post-bronchodilator FEV_1_/FVC *<* 0.7 on the diagnosis and treatment of COPD?

**Study Design and Methods:** We used a national electronic health record database to identify clinical encounters between 2007 to 2022 with patients 18 years of age and older in which a post-bronchodilator FEV_1_/FVC value was documented. An encounter was associated with a COPD diagnosis if a diagnostic code for COPD was assigned, and was associated with COPD treatment if a prescription for a medication commonly used to treat COPD was filled within 90 days. We used a regression discontinuity design to measure the effect of a post-bronchodilator FEV_1_/FVC *<* 0.7 on COPD diagnosis and treatment.

**Results:** Among 27 817 clinical encounters, involving 18 991 patients, a post-bronchodilator FEV_1_/FVC *<* 0.7 was present in 14 876 (53.4%). The presence of a documented post-bronchodilator FEV_1_/FVC *<* 0.7 had a small effect on the probability of a COPD diagnosis, increasing by 6.0% (95% confidence interval [CI] 1.1% to 10.9%) from 38.0% just above the 0.7 cutoff to 44.0% just below this cutoff. The presence of a documented post-bronchodilator FEV_1_/FVC had no effect on the probability of COPD treatment (*−*2.1%, 95% CI *−*7.2% to 3.0%).

**Interpretation:** The presence of a documented post-bronchodilator FEV_1_/FVC *<* 0.7 has only a small effect on the probability that a clinician will make a guideline-concordant diagnosis of COPD and has no effect on corresponding treatment decisions.

## Introduction

Chronic obstructive pulmonary disease (COPD) is defined by the presence of obstruction on spirometry.^1,2^ According to Global Initiative for Chronic Obstructive Lung Disease (GOLD) guidelines, obstruction is present if the post-bronchodilator forced expiratory volume in 1 second to forced vital capacity ratio (FEV_1_/FVC) is less than 0.7, with the diagnosis of COPD recommended only in patients with obstruction.^3^ Despite this, studies involving the performance of spirometry and its comparison with the prior clinical diagnosis of COPD have found that the presence of obstruction correlates only loosely with this diagnosis.^4,5^ Between 30 and 60 percent of patients who have been diagnosed with COPD do not have evidence of obstruction on spirometry,^6–10^ while between 60 and 80 percent of patients with obstruction have not been diagnosed with COPD.^11–16^

This contrast between the presence of obstruction and the diagnosis of COPD has been attributed to the underuse of spirometry,^17–21^ with the assumption that physicians would diagnose COPD in accordance with GOLD guidelines if they had access to the results of spirometry. However, while access to the results of spirometry will better position physicians to arrive at an accurate diagnosis of COPD, diagnostic accuracy depends further on the proper use of these results. Studies comparing the prior performance of spirometry with the clinical diagnosis of COPD have found that even after spirometry has been performed, the diagnosis of COPD still often fails to correspond to the recommendations of GOLD guidelines.^22–24^

To better understand the role of spirometry interpretation in medical decision making, we sought to estimate effect of the presence of a documented post-bronchodilator FEV_1_/FVC *<* 0.7 on COPD diagnosis and treatment. We hypothesized that if physicians applied GOLD guidelines to spirometry to diagnose COPD, this would yield a substantial discontinuity in the probability of a COPD diagnosis at the 0.7 cutoff; COPD would generally not be diagnosed in patients with an FEV_1_/FVC *≥* 0.7 and would generally be diagnosed in patients with an FEV_1_/FVC *<* 0.7. Because current GOLD guidelines do not recommend that spirometry directly inform treatment decisions, though treatment decisions likely follow the establishment of a COPD diagnosis, we also hypothesized that a post-bronchodilator FEV_1_/FVC *<* 0.7 would affect subsequent COPD treatment but to a lesser extent than it would affect diagnosis.

## Methods

### Data Source

We used data from the Optum Labs Data Warehouse (OLDW), a database composed of de-identified administrative claims and electronic health record (EHR) data from across the United States.^25^ EHR data included in the OLDW were derived from provider notes using a proprietary natural language processing (NLP) system.^26^ This use of NLP made it possible to link clinical data to specific clinical encounters in which physicians demonstrate access to these data by including them in their clinic notes. This linkage allowed us to study the effect of these data on diagnostic and therapeutic decision making.

### Study Population

We included clinical encounters from 2007 to 2022 in the OLDW that involved patients 18 years of age and older and that documented a post-bronchodilator FEV_1_/FVC measurement in the associated clinic note. We excluded encounters that involved only the performance of pulmonary function testing, as the diagnoses associated with these encounters are assigned prior to the encounter—and used to justify the performance of the test—rather than in response to the test results.

### Exposure and Outcomes

The exposure was the documented presence of a post-bronchodilator FEV_1_/FVC *<* 0.7. The primary outcome was whether an encounter was associated with the diagnosis of COPD. We defined an encounter as associated with a diagnosis of COPD if the encounter was assigned any ICD code for COPD, including chronic bronchitis and emphysema (**e-Table 1**). In addition to estimating the effect of the exposure on COPD diagnosis, we evaluated, as a secondary outcome, the effect of the same exposure on COPD treatment. We defined an encounter as associated with COPD treatment if an inhaler, oral corticosteroid, or other medication commonly used to treat COPD was filled within 90 days of the encounter (**e-Table 2**).

### Study Design

We used a regression discontinuity design (RDD) to estimate the effect of the exposure on the primary and secondary outcomes. An RDD is method of causal inference that allows one to estimate the effect of an exposure on an outcome when the presence of the exposure is a function of whether the value of a continuous variable falls above or below a discrete cutoff.^27,28^ Given a sufficient amount of data, one can define a small enough bandwidth around the cutoff that observations within the bandwidth that fall on on one side or the other of the cutoff will, on average, differ only by the presence or absence of the exposure of interest.^29^ The effect of the exposure can then be estimated as the discontinuity of a regression across this cutoff. In this way, an RDD can be used to provide an effect estimate from observational data that is free from unmeasured confounding.

The continuous variable in our RDD was the post-bronchodilator FEV_1_/FVC while the cutoff was the 0.7 value used by GOLD guidelines to define the presence of obstruction. The bandwidth around the 0.7 cutoff was selected for minimal coverage error with robust bias-corrected inference.^30,31^ We used local linear regression to construct the point-estimator and local quadratic regression to construct the bias correction. We followed the convention of using a triangular kernel in our RDD, so as to increase the relative weight assigned to clinical encounters with post-bronchodilator FEV_1_/FVC values within the bandwidth that were closer to the 0.7 cutoff.^30^

In addition to estimating the effect of a documented post-bronchodilator FEV_1_/FVC *<* 0.7 on the diagnosis and treatment of COPD, we also estimated its effect on specific types of COPD diagnosis—chronic bronchitis, emphysema, and other chronic obstructive lung disease—and specific classes of COPD treatment—inhaled corticosteroids, long-acting beta agonists, long-acting muscarinic antagonists, macrolide antibiotics, oral corticosteroids, phosphodiesterase-4 inhibitors, short-acting beta agonists, and short-acting muscarinic antagonists.

In our primary analysis we used a sharp RDD, as the exposure of interest was present in all encounters in which the value of the continuous variable was less than the cutoff and was absent in all encounters in which the value of the continuous variable was equal to or greater than the cutoff. In a secondary analysis we used a fuzzy RDD to assess the effect of COPD diagnosis on COPD treatment, allowing for the facts that not all patients with an FEV_1_/FVC *<* 0.7 were diagnosed with COPD and that this diagnosis could be present in patients with an FEV_1_/FVC *≥* 0.7. Much like an instrumental variable analysis, a fuzzy RDD involves two stages, the first of which associates an exogenous variable with the probability of exposure to an intervention, while the second of which associates the intervention with the outcome of interest.^32^

### Subgroups

In exploratory analyses we estimated the effect of a post-bronchodilator FEV_1_/FVC *<* 0.7 on COPD diagnosis and treatment in pre-specified subgroups defined by age *≥* 65, gender, race, history of tobacco use, history of COPD, encounter type, and physician specialty.

### Validity Assessment and Sensitivity Analysis

We performed multiple tests to assess the validity of our RDD and further assessed the sensitivity of our results to different modeling assumptions. These analyses are described in our supplementary methods.

### Statistical Analysis

All statistical tests were two sided and a *P* value *<* 0.05 was interpreted as statistically significant. R version 4.2.1 was used for data analysis.^33^ The rdrobust package was used to perform the RDD.^34^ We followed the STROBE checklist for reporting observational studies in epidemiology (**e-Table 3**).^35^ As the study involved the use of secondary, de-identified data it did not represent human subjects research and Institutional Review Board approval was not necessary.

## Results

We identified 27 817 clinical encounters in which a post-bronchodilator FEV_1_/FVC measurement was documented, involving 18 991 different patients and more than 2 038 different physicians (**Table 1**). The encounters most often involved women (*N* = 14 517, 52%), non-Hispanic White patients (*N* = 25 083, 90%), patients at least 65 years old (*N* = 14 586, 52%), and patients with a history of tobacco use (*N* = 14 909, 54%). A total of 6 674 (24.0%) encounters involved patients with a prior COPD diagnosis. The primary physician specialty was documented for 11 755 encounters, with 1 932 (6.9%) encounters with pulmonologists, 6 705 (24.1%) with internists or non-pulmonologist internal medicine subspecialties, 643 (2.3%) with family medicine or general practitioners, 1 527 (5.5%) with emergency medicine physicians, and 948 (3.4%) with surgeons.

**Table 1.** Encounter Characteristics.

|  | All<br>(N = 27 817) | FEV<sub>1</sub>/FVC < 0.7<br>(N = 14 876) | FEV<sub>1</sub>/FVC ≥ 0.7<br>(N = 12 941) |
| --- | --- | --- | --- |
| **Patient Age** |  |  |  |
| 18 - 40 years | 2 577 (9.3%) | 754 (29.3%) | 1 823 (70.7%) |
| 41 - 64 years | 10 654 (38.3%) | 5 278 (49.5%) | 5 376 (50.5%) |
| 65 - 90 years | 14 586 (52.4%) | 8 844 (60.6%) | 5 742 (39.4%) |
| **Patient Gender** |  |  |  |
| Men | 13 295 (47.8%) | 7 812 (58.8%) | 5 483 (41.2%) |
| Women | 14 517 (52.2%) | 7 060 (48.6%) | 7 457 (51.4%) |
| **Patient Race and Ethnicity** |  |  |  |
| Asian | 134 (0.5%) | 37 (27.6%) | 97 (72.4%) |
| Hispanic | 531 (1.9%) | 165 (31.1%) | 366 (68.9%) |
| Non-Hispanic Black | 1 252 (4.5%) | 535 (42.7%) | 717 (57.3%) |
| Non-Hispanic White | 25 083 (90.2%) | 13 736 (54.8%) | 11 347 (45.2%) |
| Other | 817 (2.9%) | 403 (49.3%) | 414 (50.7%) |
| **Patient History of Tobacco Use** |  |  |  |
| Yes | 14 909 (53.6%) | 9 451 (63.4%) | 5 458 (36.6%) |
| No | 12 908 (46.4%) | 5 425 (42.0%) | 7 483 (58.0%) |
| **Patient History of COPD Diagnosis** |  |  |  |
| Yes | 6 674 (24.0%) | 5 065 (75.9%) | 1 609 (24.1%) |
| No | 21 143 (76.0%) | 9 811 (46.4%) | 11 332 (53.6%) |
| **Encounter Type** |  |  |  |
| Inpatient | 10 680 (38.4%) | 5 868 (54.9%) | 4 812 (45.1%) |
| Outpatient | 17 137 (61.6%) | 9 008 (52.6%) | 8 129 (47.4%) |
| **Physician Specialty** |  |  |  |
| Pulmonology | 1 932 (6.9%) | 1 120 (58.0%) | 812 (42.0%) |
| Other Internal Medicine | 6 705 (24.1%) | 3 644 (54.3%) | 3 061 (45.7%) |
| Family Medicine | 643 (2.3%) | 319 (49.6%) | 324 (50.4%) |
| Emergency Medicine | 1 527 (5.5%) | 910 (59.6%) | 617 (40.4%) |
| Surgery | 948 (3.4%) | 438 (46.2%) | 510 (53.8%) |
| Unknown | 16 062 (57.7%) | 8 445 (52.6%) | 7 617 (47.4%) |

COPD = chronic obstructive pulmonary disease; FEV<sub>1</sub> = forced expiratory volume in 1 second; FVC = forced vital capacity.

COPD was diagnosed in 12 697 (45.6%) encounters, including 3 219 (25.3%) encounters in which a post-bronchodilator FEV_1_/FVC was *≥* 0.7. COPD was not diagnosed in 15 120 (54.4%) encounters, including 5 398 (35.7%) encounters in which a post-bronchodilator FEV_1_/FVC was *<* 0.7. A total of 16 515 (59.4%) encounters were associated with COPD treatment, including 6 608 (40.0%) in which a post-bronchodilator FEV_1_/FVC was *≥* 0.7. A total of 11 302 (40.6%) encounters were not associated with COPD treatment, including 4 969 (44.0%) encounters in which a post-bronchodilator FEV_1_/FVC was *<* 0.7.

The presence of a documented post-bronchodilator FEV_1_/FVC *<* 0.7 had a small but statistically significant effect on the diagnosis of COPD (**Figure 1**). The probability of a COPD diagnosis increased from 38.0% just above the 0.7 post-bronchodilator FEV_1_/FVC cutoff to 44.0% just below this cutoff, a discontinuity of 6.0% (95% CI 1.1% to 10.9%, *P* value = 0.016) at the cutoff (**Table 2**). This effect was not seen with pre-bronchodilator spirometry (**e-Table 4**) and was seen only in the diagnosis of chronic obstruction (5.4% 95%CI 0.9% to 9.8%) and not in the diagnosis of chronic bronchitis (1.3%, 95% CI *−*2.2% to 4.8%) or emphysema (*−*0.2%, 95% CI *−*2.0% to 1.7%) (**e-Table 5**).

**Table 2.** Effect of a Post-Bronchodilator FEV_1_/FVC *<* 0.7 on COPD Diagnosis and Treatment.

| Outcome | Probability of Outcome |  | Change in Probability<br>at 0.7 Cutoff (95% CI)<sup>a</sup> |
| --- | --- | --- | --- |
|  | Above<br>0.7 Cutoff | Below<br>0.7 Cutoff |  |
| COPD Diagnosis<sup>b</sup> | 38.0% | 44.0% | 6.0% (1.1% to 10.9%) |
| COPD Treatment<sup>c</sup> | 60.7% | 58.7% | −2.1% (−7.2% to 3.0%) |

CI = confidence interval; COPD = chronic obstructive pulmonary disease; $FEV_1$ = forced expiratory volume in 1 second; FVC = forced vital capacity.
<sup>a</sup> Bias-corrected discontinuity estimate with data-driven bandwidth selection and robust standard errors.
<sup>b</sup> A clinical encounter is associated with a COPD diagnosis if the encounter is assigned an ICD code for COPD, including chronic bronchitis and emphysema.
<sup>c</sup> A clinical encounter is associated with treatment for COPD if a medication used to treat COPD is prescribed within 90 days following the encounter.

**Table 3.** Effect of a Post-Bronchodilator FEV_1_/FVC *<* 0.7 on COPD Diagnosis and Treatment by Subgroup.

|  | Probability of COPD Diagnosis<sup>a</sup> |  |  | Probability of COPD Treatment<sup>b</sup> |  |  |
| --- | --- | --- | --- | --- | --- | --- |
|  | Above<br>0.7 Cutoff | Below<br>0.7 Cutoff | Change in Probability<br>at 0.7 Cutoff (95% CI)<sup>c</sup> | Above<br>0.7 Cutoff | Below<br>0.7 Cutoff | Change in Probability<br>at 0.7 Cutoff (95% CI)<sup>c</sup> |
| **Age** |  |  |  |  |  |  |
| < 65 years | 36.7% | 41.6% | 4.9% (−1.6% to 11.3%) | 63.8% | 61.3% | −2.5% (−9.0% to 4.1%) |
| ≥ 65 years | 39.3% | 46.9% | 7.6% (0.6% to 14.6%) | 58.4% | 56.1% | −2.2% (−9.1% to 4.6%) |
| **Gender** |  |  |  |  |  |  |
| Men | 38.4% | 45.7% | 7.3% (0.3% to 14.2%) | 59.9% | 53.1% | −6.7% (−14.5% to 1.0%) |
| Women | 37.8% | 42.7% | 4.8% (−1.6% to 11.3%) | 61.6% | 64.6% | 3.0% (−2.4% to 8.5%) |
| **Race** |  |  |  |  |  |  |
| White | 38.4% | 44.6% | 6.2% (1.1% to 11.3%) | 60.3% | 58.2% | −2.1% (−7.4% to 3.2%) |
| Black | 37.1% | 32.0% | −5.2% (−21.9% to 11.5%) | 68.1% | 66.8% | −1.3% (−24.1% to 21.5%) |
| **History of Tobacco Use** |  |  |  |  |  |  |
| Yes | 54.6% | 59.9% | 5.3% (−1.6% to 12.2%) | 62.8% | 58.7% | −4.1% (−10.4% to 2.3%) |
| No | 23.0% | 26.7% | 3.7% (−2.5% to 9.9%) | 60.0% | 56.7% | −3.3% (−11.1% to 4.5%) |
| **History of COPD** |  |  |  |  |  |  |
| Yes | 84.8% | 85.8% | 0.9% (−6.1% to 7.9%) | 60.7% | 65.0% | 4.3% (−5.6% to 14.2%) |
| No | 26.1% | 32.7% | 6.6% (1.4% to 11.7%) | 61.0% | 57.1% | −3.8% (−9.8% to 2.2%) |
| **Encounter Type** |  |  |  |  |  |  |
| Inpatient | 47.4% | 55.3% | 7.9% (−0.4% to 16.2%) | 55.5% | 55.2% | −0.4% (−8.7% to 8.0%) |
| Outpatient | 32.3% | 37.8% | 5.5% (−0.1% to 11.1%) | 63.7% | 61.1% | −2.6% (−8.6% to 3.3%) |
| **Physician Specialty** |  |  |  |  |  |  |
| Pulmonology | 29.9% | 39.7% | 9.8% (−4.9% to 24.4%) | 66.5% | 71.0% | 4.5% (−16.1% to 25.0%) |
| Other Internal Medicine | 51.5% | 55.9% | 4.5% (−3.8% to 12.7%) | 58.2% | 59.4% | 1.2% (−8.2% to 10.6%) |
| Family Medicine | 27.5% | 49.4% | 22.0% (−5.6% to 49.5%) | 63.4% | 65.8% | 2.4% (−35.2% to 40.0%) |
| Emergency Medicine | 43.7% | 73.0% | 29.3% (7.3% to 51.3%) | 64.0% | 65.1% | 1.1% (−17.4% to 19.5%) |
| Surgery | 30.1% | 15.4% | −14.8% (−36.1% to 6.6%) | 25.8% | 26.3% | 0.5% (−22.9% to 23.8%) |

CI = confidence interval; COPD = chronic obstructive pulmonary disease.
<sup>a</sup> A clinical encounter is associated with a COPD diagnosis if the encounter is assigned an ICD code for COPD, including chronic bronchitis and emphysema.
<sup>b</sup> A clinical encounter is associated with treatment for COPD if a medication used to treat COPD is prescribed within 90 days following the encounter.
<sup>c</sup> Bias-corrected discontinuity estimate with data-driven bandwidth selection and robust standard errors.

**Figure 1.**
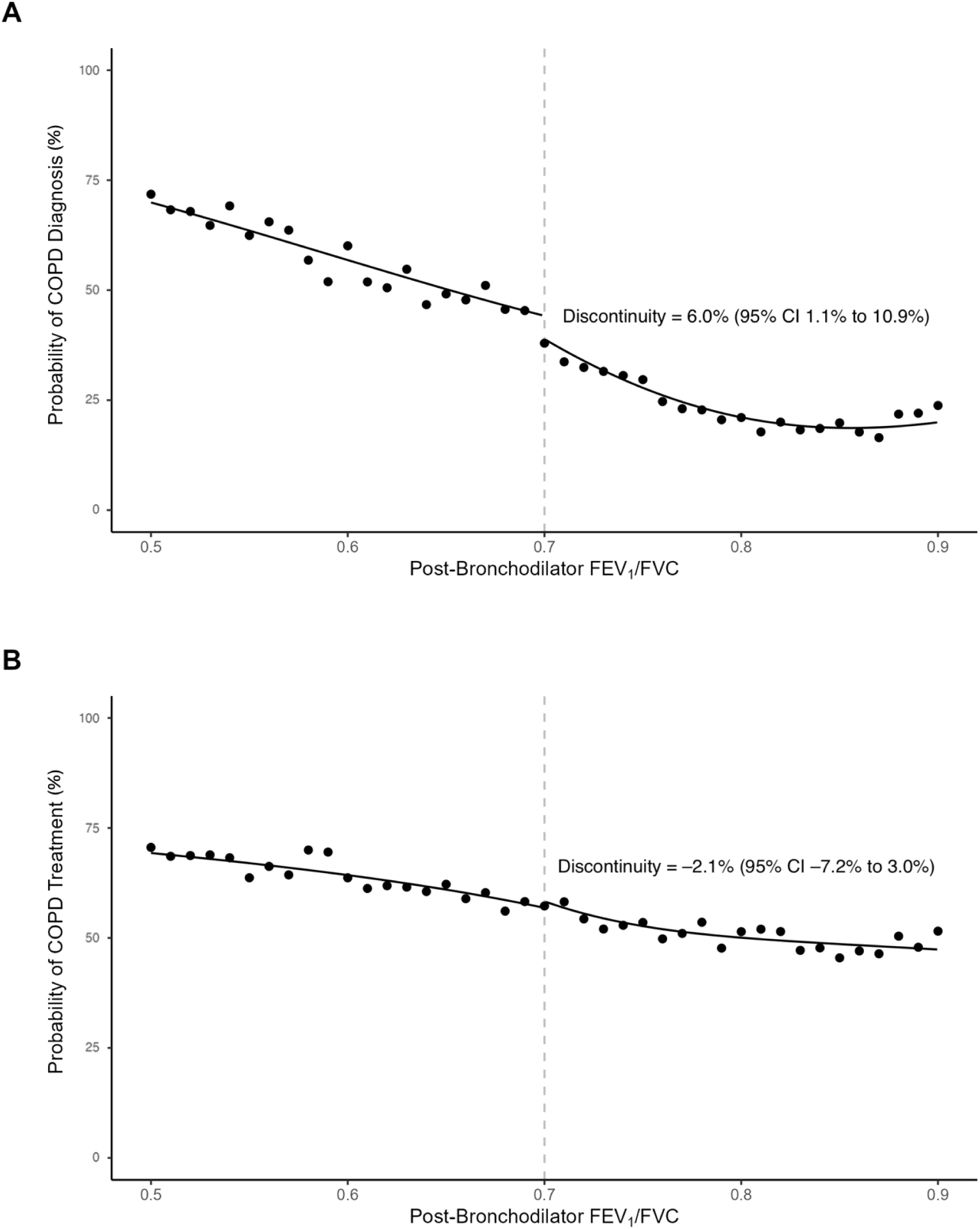
Association between Post-Bronchodilator FEV_1_/FVC and the Diagnosis and Treatment of COPD. Binscatter plots depict the association between post-bronchodilator FEV_1_/FVC values and the probability that COPD is (A) diagnosed and (B) treated. Post-bronchodilator FEV_1_/FVC values are binned at the 0.01 interval. The vertical dashed line represents the post-bronchodilator FEV_1_/FVC cutoff of 0.7 recommended by GOLD guidelines. Abbreviations: COPD = chronic obstructive pulmonary disease; FEV_1_ = forced expiratory volume in 1 second; FVC = forced vital capacity; GOLD = Global Initiative for Chronic Obstructive Lung Disease.

The presence of a post-bronchodilator FEV_1_/FVC *<* 0.7 did not affect COPD treatment (**Figure 1**). The probability of treatment was 60.7% just above the 0.7 cutoff and 58.7% just below this cutoff, with a discontinuity of *−*2.1% (95% CI *−*7.2% to 3.0%) at the cutoff (**Table 2**). A significant effect was seen by treatment type only in the case of roflumilast (0.9%, 95% CI 0.1% to 1.7%) (**e-Table 6**). In our secondary analysis applying a fuzzy RDD to assess the effect of a COPD diagnosis on COPD treatment, the diagnosis of COPD did not have a significant effect on COPD treatment (48.1%, 95% CI *−*55.7% to 152.0%).

### Subgroup Analysis

Our exploratory subgroup analysis suggested that physician speciality and the history of a COPD diagnosis may both impact the role of spirometry in COPD diagnosis. While the presence of a post-bronchodilator FEV_1_/FVC *<* 0.7 did not have a significant effect on the diagnosis of COPD among pulmonologists (9.8%, 95% CI *−*4.9% to 24.4%) or other internal medicine physicians (4.5%, 95% CI *−*3.8% to 12.7%), it did increase the probability of a COPD diagnosis in encounters with emergency medicine physicians (29.3%, 95% CI 7.3% to 51.3%). Notably, in patients with a prior diagnosis of COPD, the presence of a post-bronchodilator FEV_1_/FVC *<* 0.7 had no effect on the diagnosis of COPD (0.9%, 95% CI *−*6.1% to 7.9%), with a probability of diagnosis of 84.8% above the cutoff and a probability of 85.8% below the cutoff.

### Validity Assessment and Sensitivity Analysis

As detailed in our supplementary results, the validity of our RDD was supported by the absence of evidence of manipulation of the continuous variable along with the negative results of all placebo tests,^29,36^ while our sensitivity analysis demonstrated that our results were generally robust to the adoption of different modeling assumptions.

## Discussion

We applied an RDD to a national EHR database and found that the presence of a documented post-bronchodilator FEV_1_/FVC *<* 0.7 only slightly increased the probability of a diagnosis of COPD. In the same RDD, the cutoff had no effect on COPD treatment even across a range of clinical contexts and encounter types. These findings suggest that GOLD guidelines have less of an effect on clinical decision making than has been assumed and that the performance of spirometry is insufficient to guarantee the diagnosis of COPD in accordance with these guidelines.

There are multiple potential etiologies for the observed discrepancy between the recommendations of GOLD guidelines and the diagnosis of COPD in this study. Though the OLDW database uses NLP to extract data from clinic notes, spirometry data may have been auto-populated and physicians who included these data in their documentation may nonetheless have been unaware of them. Other physicians may have been aware of the results of spirometry, but unaware of their implications. As European Respiratory Society and American Thoracic Society (ERS/ATS) guidelines for spirometry interpretation do not provide physicians with recommendations regarding the diagnostic implications of spirometry, physicians must independently decide if test results are consistent with a diagnosis of COPD.^37^ Other physicians may have relied on different spirometric criteria to determine the presence of obstruction. There is a lack of consensus regarding the use of the fixed 0.7 cutoff recommended by GOLD to define the presence of obstruction on spirometry and some physicians may have instead used the FEV_1_/FVC lower limit of normal to identify obstruction.^37^ Finally, some physicians may have been aware of the diagnostic implications of spirometry and yet set aside the 0.7 cutoff as too simple a tool to apply to a clinically heterogenous disease such as COPD.^38,39^

If physicians are largely unaware of the diagnostic implications of a post-bronchodilator FEV_1_/FVC *<* 0.7, our study suggests a role for the use of clinical decision support to help physicians diagnose COPD in a manner concordant with GOLD guidelines.^40^ Indeed, if this is the case, our study suggests that in the absence of such decision support, COPD misdiagnosis may persist with some frequency even after spirometry has been performed. Attempts to improve COPD diagnosis, simply by increasing the performance of spirometry, as with recent proposals to screen for COPD with spirometry, will be less successful than imagined if spirometry interpretation—rather than just spirometry performance—represents an important limiting factor in COPD diagnosis.^41^

On the other hand, if the discrepancy between the recommendations of GOLD guidelines and the clinical diagnosis of COPD stems from the decision on the part of clinicians to depart from these guidelines, then simply alerting physicians to the fact that the post-bronchodilator FEV_1_/FVC is less than 0.7 will have little effect on clinical practice. If this is the case, COPD diagnosis might be advanced by replacing the simple 0.7 cutoff with a more robust model of airway obstruction.^42,43^

We found that the presence of a post-bronchodilator FEV_1_/FVC *<* 0.7 had no effect on COPD treatment. While GOLD had previously recommended the use of spirometry to directly inform treatment decisions, more recent guidelines recommend instead that treatment decisions be informed by exacerbation history and respiratory symptoms. As such, the presence of a post-bronchodilator FEV_1_/FVC *<* 0.7 was expected to have had less of an effect on COPD treatment than on diagnosis.^3^ Nonetheless, as GOLD guidelines have consistently recommended at minimum the prescription of a short-acting bronchodilator for patients with COPD, the presence of a post-bronchodilator FEV_1_/FVC *<* 0.7 would be expected to have at least some effect on COPD treatment.^44^

The results of our exploratory subgroup analysis suggest that physicians with different medical specialties may use spirometry in different ways to diagnose COPD. While the presence of a post-bronchodilator FEV_1_/FVC *<* 0.7 had a significant effect on the diagnosis of COPD by emergency medicine physicians, a similar effect was not seen among other type of physicians. This finding suggests that physicians who have a longitudinal relationship with their patients may rely more on history and symptoms to diagnose COPD while physicians without this type of clinical relationship may rely more on objective data in the form of spirometry. Likewise, while the presence of a post-bronchodilator FEV_1_/FVC *<* 0.7 had a significant effect on the diagnosis of COPD in patients who had not been previously been diagnosed, it had no effect on the diagnosis of patients who already carried this diagnosis. Diagnostic momentum appears to play a significant role in COPD diagnosis and once a diagnosis of COPD has been made, spirometry seems to have little effect upon it.

Finally, this study provides the first estimate of the effect of spirometry interpretation on diagnostic and therapeutic decision making in clinical practice. While several recent studies have speculated as to the downstream clinical consequences that follow from the recommended adoption of race-neutral reference equations, these consequences have yet to be studied empirically.^45–49^ Our finding that the presence of a post-bronchodilator FEV_1_/FVC *<* 0.7 has only a minimal effect on COPD diagnosis and no effect on COPD treatment challenges the assumption that whether a spirometric parameter falls above or below a lower limit of normal—the effect of adopting one set of reference equations or another—results in corresponding changes in clinical decision making. The relationship between spirometry interpretation and such clinical decisions is not as straightforward as has been assumed and empirical studies are needed to estimate the clinical consequences that follow from the adoption of novel reference equations.

This study has several strengths. First, we used a national EHR database and included encounters involving tens of thousands of patients and thousands of physicians. Second, the use of NLP to extract spirometry data from clinic notes allowed us to link these data to specific clinical encounters and conclude not only that spirometry had been performed but that the results of such performance were documented and thus accessible to physicians. Third, our use of an RDD mitigated the impact of unmeasured confounding on our effect estimates. Fourth, we performed an extensive sensitivity analysis and found that our results were generally insensitive to different modeling assumptions.

This study also has several limitations. First, we were unable to identify the FEV_1_ percent predicted values or FEV_1_/FVC lower limit of normal values to which the physicians in our cohort had access, and we were thus unable to assess the effect these components of spirometry may have had on diagnostic and therapeutic decision making. Second, though our data were drawn from a national EHR database, post-bronchodilator spirometry data were available for only a subset of the patients in this database. The selection effects mediating the inclusion of these data are unknown and may limit the external validity of our findings. Third, our use of ICD codes to associate COPD diagnoses with clinical encounters likely underestimates the prevalence of COPD diagnosis in our cohort as it is unlikely that an ICD code for COPD will be assigned to each clinical encounter with a patient diagnosed with COPD, given that some encounters will involve medical issues unrelated to this diagnosis. Fourth, our use of prescription data to associate COPD treatment with a clinical encounter likely overestimates the prevalence of COPD treatment as we were unable to identify the specific rationale for each prescription and many of these medications can be used to treat other diseases as well. Fifth, a fundamental limitation of the RDD is that it provides a local estimate of an effect at a cutoff. In our analysis we were thus unable to estimate the effect of a post-bronchodilator FEV_1_/FVC *<* 0.7 for clinical encounters with FEV_1_/FVC values that are far from the 0.7 cutoff.^50^

## Interpretation

In conclusion, we found that the presence of a documented post-bronchodilator FEV_1_/FVC *<* 0.7 had only a small effect on the diagnosis of COPD and had no effect on COPD treatment. These findings suggest that the prevailing, guideline-recommended diagnostic cutoff for COPD may not meaningfully affect clinical decision making. Further work is needed to accurately and reliably incorporate spirometry data into the diagnostic process for COPD.

## Data Availability

All data produced in the present study are available upon reasonable request to the authors.

