## Supplement for "The Effect of a Post-Bronchodilator FEV_1_/FVC *<* 0.7 on COPD Diagnosis and Treatment: A Regression Discontinuity Design"

**Supplementary Methods**

**Supplementary Results**

<u>Tables</u>

**e-Table 1** ICD Codes for Chronic Obstructive Pulmonary Disease Diagnosis

**e-Table 2** Medications for Chronic Obstructive Pulmonary Disease Treatment

**e-Table 3** STROBE Checklist

**e-Table 4** Effect of Pre- and Post-Bronchodilator $FEV_1/FVC < 0.7$ on COPD Diagnosis

**e-Table 5** Effect of $FEV_1/FVC$ Cutoff on COPD Diagnosis by Diagnosis Type

**e-Table 6** Effect of $FEV_1/FVC$ Cutoff on COPD Treatment by Treatment Type

**e-Table 7** Conventional and Bias-Corrected Effect Estimates

**e-Table 8** Adjusted and Unadjusted Effect Estimates

**e-Table 9** Kernel Shape and Effect Estimates

**e-Table 10** Polynomial Order and Effect Estimates

**e-Table 11** Bandwidth Selection Methods and Effect Estimates

**e-Table 12** Variance-Covariance Matrix and Effect Estimates

<u>Figures</u>

**e-Figure 1** CONSORT Diagram

**e-Figure 2** Distribution of Post-Bronchodilator $FEV_1/FVC$ Values

**e-Figure 3** Distribution of Select Covariates at the Cutoff

**e-Figure 4** Association between Cutoff and Effect Estimates

**e-Figure 5** Association between Main Bandwidth and Effect Estimates

**e-Figure 6** Association between Bias Bandwidth and Effect Estimates

**e-Figure 7** Sensitivity of Effect Estimate to Time to Diagnosis

**e-Figure 8** Sensitivity of Effect Estimate to Time to Treatment

### Supplementary Methods

#### Validity Assessment

We performed multiple tests to assess the validity of the RDD. First, we tested for differences in the density of the post-bronchodilator $FEV_1/FVC$ values around the 0.7 cutoff to assess for potential manipulation of the continuous variable. Manipulation can occur when the value of the continuous variable is changed in response to whether it falls above or below the cutoff, with the presence of manipulation invalidating the assumption that observations within the bandwidth falling above or below the cutoff will be otherwise identical. Second, we performed placebo tests to test the effect of a post-bronchodilator $FEV_1/FVC < 0.7$ on age, sex, race, tobacco use, history of COPD, and clinical encounter type, to assess covariate balance. Third, we compared the effect estimates using a 0.7 cutoff to estimates produced using all cutoff values at a 0.01 interval between 0.5 and 0.9. Fourth, we compared effect estimates using pre- and post-bronchodilator $FEV_1/FVC$ values, with the expectation that an effect would be greater in the latter.

#### Sensitivity Analysis

We assessed the sensitivity of our results to different modeling assumptions. First, we compared the use of bias-corrected estimates and robust standard errors to the use of conventional estimates and conventional standard errors. Second, we compared an unadjusted RDD to an RDD adjusted for age, sex, race, ethnicity, tobacco use, history of COPD diagnosis, encounter type, and physician specialty. Third, we compared the use of different kernel shapes in our RDD. Fourth, we compared the use of different orders for the polynomials used to construct the point estimator and bias-correction estimator in our RDD. Fifth, we compared the use of different methods for bandwidth selection. Sixth, we compared the use of different bandwidth values to construct the point estimator and bias-correction estimator. Seventh, we compared the use of different procedures to compute the variance-covariance matrix estimator. Eighth, we compared the use of different cutoff dates for the association of COPD diagnosis and treatment with clinical encounters, ranging from the day of the encounter to 365 days after the encounter.

### Supplementary Results

#### Validity Assessment

The validity of the RDD was supported by the absence of evidence of manipulation of the continuous variable ($P = 0.186$, **e-Figure 2**) and by the negative results of all placebo tests (**e-Figure 3**). Assessing the effect of different post-bronchodilator $FEV_1$/FVC cutoff values on COPD diagnosis we found that cutoffs of 0.58 and 0.67 were associated with a significant increase in the probability of diagnosis, while cutoffs of 0.60, 0.66, and 0.67 were associated with a significant decrease in the probability of diagnosis (**e-Figure 4**). These results likely reflect Type I errors as these cutoffs are not expected to have an effect on COPD diagnosis and the directionality of these effects were roughly even.

#### Sensitivity Analysis

Our results were generally robust to the adoption of different modeling assumptions. The presence of a post-bronchodilator $FEV_1/FVC < 0.7$ was found to have an effect on COPD diagnosis but not treatment with conventional estimates and conventional standard errors (**e-Table 7**), with an adjusted analysis (**e-Table 8**), with a rectangular kernel (**e-Table 9**), with polynomial orders for the point estimator of less than 2 and for the bias-correction estimator of less than 4 (**e-Table 10**), with most approaches to bandwidth selection (**e-Table 11**), with different procedures to compute the variance-covariance matrix estimator (**e-Table 12**), with different main (**e-Figure 5**) and bias (**e-Figure 6**) bandwidths, and with different time periods used to associate diagnosis (**e-Figure 7**) and treatment (**e-Figure 8**) with an encounter.

**e-Table 1.** ICD Codes for Chronic Obstructive Pulmonary Disease Diagnosis

| | **Code** | **Diagnosis** |
|---|---|---|
| ICD-9 | 491 | Chronic Bronchitis |
| | 491.0 | Simple Chronic Bronchitis |
| | 491.1 | Mucopurulent Chronic Bronchitis |
| | 491.2 | Obstructive Chronic Bronchitis |
| | 491.20 | Obstructive Chronic Bronchitis without Exacerbation |
| | 491.21 | Obstructive Chronic Bronchitis with Exacerbation |
| | 491.22 | Obstructive Chronic Bronchitis with Acute Bronchitis |
| | 491.8 | Other Chronic Bronchitis |
| | 491.9 | Unspecified Chronic Bronchitis |
| | 492 | Emphysema |
| | 492.0 | Emphysematous Bleb |
| | 492.8 | Other Emphysema |
| | 496 | Chronic Airway Obstruction, Not Elsewhere Classified |
| ICD-10 | J41 | Simple and Mucopurulent Chronic Bronchitis |
| | J41.0 | Simple Chronic Bronchitis |
| | J41.1 | Mucopurulent Chronic Bronchitis |
| | J41.2 | Mixed Simple and Mucopurulent Chronic Bronchitis |
| | J42 | Unspecified Chronic Bronchitis |
| | J43 | Emphysema |
| | J43.0 | Unilateral Emphysema |
| | J43.1 | Panlobular Emphysema |
| | J43.2 | Centrilobular Emphysema |
| | J43.8 | Other Emphysema |
| | J43.9 | Emphysema, Unspecified |
| | J44 | Other Chronic Obstructive Pulmonary Disease |
| | J44.0 | Chronic Obstructive Pulmonary Disease with Lower Respiratory infection |
| | J44.1 | Chronic Obstructive Pulmonary Disease with Exacerbation |
| | J44.8 | Other Specified Chronic Obstructive Pulmonary Disease |
| | J44.89 | Other Specified Chronic Obstructive Pulmonary Disease |
| | J44.9 | Chronic Obstructive Pulmonary Disease, Unspecified |

Abbreviations: ICD = International Classification of Diseases.

**e-Table 2.** Medications for Chronic Obstructive Pulmonary Disease Treatment

| **Class** | **Medication** |
|---|---|
| ICS | Beclomethasone<br>Budesonide<br>Fluticasone<br>Mometasone<br>Triamcinolone |
| LABA | Arformoterol<br>Formoterol<br>Indacaterol<br>Oldaterol<br>Salmeterol<br>Vilanterol |
| LAMA | Aclidinium<br>Glycopyrrolate<br>Tiotropium<br>Umeclidinium |
| Macrolide | Azithromycin |
| OCS | Methylprednisolone<br>Dexamethasone<br>Prednisone |
| PDE4 Inhibitor | Roflumilast |
| SABA | Albuterol<br>Bitolterol<br>Levalbuterol<br>Pirbuterol<br>Terbutaline |
| SAMA | Ipratropium |

Abbreviations: ICS = inhaled corticosteroid; LABA = long-acting beta agonist; LAMA = long-acting muscarinic antagonist; OCS = oral corticosteroid; PDE4 = phosphodiesterase-4; SABA = short-acting beta agonist; SAMA = short-acting muscarinic antagonist.

**e-Table 3.** STROBE Checklist

| | Item | Recommendation | Page |
|---|---|---|---|
| Title and Abstract | 1a | Indicate study design | 2 |
| | 1b | Informative and balanced summary | 2 |
| *Introduction* | | | |
| Background | 2 | Explain background and rationale | 4 |
| Objectives | 3 | State specific objectives | 4 |
| *Methods* | | | |
| Study Design | 4 | Present key elements | 7 |
| Setting | 5 | Describe setting, locations, and relevant dates | 6 |
| Participants | 6 | Give eligibility criteria and selection | 6 |
| Variables | 7 | Define outcomes, exposures, and predictors | 6 |
| Data Sources | 8 | Give source and assessment of each variable | 6 |
| Bias | 9 | Describe efforts to address bias | 8 |
| Study Size | 10 | Explain how the study size was determined | 6 |
| Quantitative Variables | 11 | Explain handling of quantitative variables | 6 |
| Statistical Methods | 12a | Describe all statistical methods | 7 |
| | 12b | Describe methods for subgroups and interactions | 8 |
| | 12c | Explain how missing data were addressed | 7 |
| | 12d | Describe sampling strategy | 6 |
| | 12e | Describe sensitivity analysis | S2 |
| *Results* | | | |
| Participants | 13a | Report number of individuals at each stage of study | S16 |
| | 13b | Give reasons for non-participation at each stage | S16 |
| | 13c | Consider use of a flow diagram | S16 |
| Descriptive Data | 14a | Give characteristics of study participants | 24 |
| | 14b | Indicate number of participants with missing data | S16 |
| Outcome Data | 15 | Report number of outcome events | 24 |
| Main Results | 16a | Give adjusted and unadjusted estimates | S11 |
| | 16b | Report category boundaries | N/A |
| | 16c | Translate relative risk into absolute risk | N/A |
| Other Analyses | 17 | Report other analyses performed | 11 |
| *Discussion* | | | |
| Key Results | 18 | Summarize key results | 12 |
| Limitations | 19 | Discuss limitations of the study | 14 |
| Interpretation | 20 | Give an overall interpretation | 16 |
| Generalizability | 21 | Discuss external validity | 15 |
| *Other Information* | | | |
| Funding | 22 | Give source of funding | 0 |

**e-Table 4.** Effect of Pre- and Post-Bronchodilator $FEV_1/FVC < 0.7$ on COPD Diagnosis

| | **Change in Probability of COPD Diagnosis at 0.7 Cutoff (95% CI)**[a] |
|---|---|
| Pre-Bronchodilator Spirometry | −2.8% (−3.6% to −1.9%) |
| Post-Bronchodilator Spirometry | 6.0% (1.1% to 10.9%) |

Abbreviations: CI = confidence interval; COPD = chronic obstructive pulmonary disease; $FEV_1$ = forced expiratory volume in 1 second; FVC = forced vital capacity.

[a] Bias-corrected discontinuity estimate with data-driven bandwidth selection and robust standard errors.

**e-Table 5.** Effect of a Post-Bronchodilator $FEV_1/FVC < 0.7$ on COPD Diagnosis by Diagnosis Type

| | **Probability of COPD Diagnosis** | | |
|---|---|---|---|
| **Diagnosis** | **Above 0.7 Cutoff** | **Below 0.7 Cutoff** | **Change in Probability at 0.7 Cutoff (95% CI)**[a] |
| Chronic Bronchitis | 13.5% | 14.8% | 1.3% (−2.2% to 4.8%) |
| Emphysema | 4.8% | 4.6% | −0.2% (−2.0% to 1.7%) |
| Other Chronic Obstruction | 29.1% | 34.5% | 5.4% (0.9% to 9.8%) |

Abbreviations: CI = confidence interval; COPD = chronic obstructive pulmonary disease; GOLD = Global Initiative for Chronic Obstructive Lung Disease.

[a] Bias-corrected discontinuity estimate with data-driven bandwidth selection and robust standard errors.

**e-Table 6.** Effect of a Post-Bronchodilator $FEV_1/FVC < 0.7$ on COPD Treatment by Treatment Type

| | **Probability of COPD Treatment** | | |
|---|---|---|---|
| **Treatment** | **Above 0.7 Cutoff** | **Below 0.7 Cutoff** | **Change in Probability at 0.7 Cutoff (95% CI)**[a] |
| Azithromycin | 8.0% | 7.5% | −0.5% (−3.1% to 2.0%) |
| ICS | 27.1% | 30.4% | 3.3% (−1.0% to 7.7%) |
| LABA | 22.3% | 26.1% | 3.8% (−0.2% to 7.9%) |
| LAMA | 11.8% | 10.7% | −1.1% (−4.3% to 2.1%) |
| OCS | 28.2% | 24.7% | −3.5% (−8.0% to 1.0%) |
| Roflumilast | 0.1% | 1.0% | 0.9% (0.1% to 1.7%) |
| SABA | 32.1% | 31.9% | −0.1% (−5.0% to 4.8%) |
| SAMA | 10.5% | 9.3% | −1.2% (−4.3% to 1.9%) |

Abbreviations: CI = confidence interval; COPD = chronic obstructive pulmonary disease; ICS = inhaled corticosteroid; LABA = long-acting beta agonist; LAMA = long-acting muscarinic antagonist; OCS = oral corticosteroid; SABA = short-acting beta agonist; SAMA = short-acting muscarinic antagonist.

[a] Bias-corrected discontinuity estimate with data-driven bandwidth selection and robust standard errors.

**e-Table 7.** Conventional and Bias-Corrected Effect Estimates

| | **COPD Diagnosis** | **COPD Treatment** |
|---|---|---|
| **Estimate** | **Effect (95% CI)[a]** | **Effect (95% CI)[a]** |
| Conventional Estimate with Conventional Standard Errors | 6.5% (2.3% to 10.6%) | −1.4% (−5.8% to 2.9%) |
| Bias-Corrected Estimate with Conventional Standard Errors | 6.0% (1.9% to 10.1%) | −2.1% (−6.4% to 2.3%) |
| Bias-Corrected Estimate with Robust Standard Errors | 6.0% (1.1% to 10.9%) | −2.1% (−7.2% to 3.0%) |

Abbreviations: CI = confidence interval; COPD = chronic obstructive pulmonary disease.

[a] Bias-corrected discontinuity estimate with data-driven bandwidth selection and robust standard errors.

**e-Table 8.** Adjusted and Unadjusted Effect Estimates

| | **COPD Diagnosis** | **COPD Treatment** |
| --- | --- | --- |
| | **Effect (95% CI)**[a] | **Effect (95% CI)**[a] |
| Adjusted[b] | 4.6% (0.5% to 8.7%) | −2.8% (−7.7% to 2.2%) |
| Unadjusted | 6.0% (1.1% to 10.9%) | −2.1% (−7.2% to 3.0%) |

Abbreviations: CI = confidence interval; COPD = chronic obstructive pulmonary disease.

[a] Bias-corrected discontinuity estimate with data-driven bandwidth selection and robust standard errors.

[b] Adjusted for patient age, gender, race, ethnicity, history of tobacco use, history of COPD diagnosis, encounter type, and physician specialty.

**e-Table 9.** Kernel Shape and Effect Estimates

| | COPD Diagnosis | COPD Treatment |
|---|---|---|
| **Kernel Shape** | **Effect (95% CI)**[a] | **Effect (95% CI)**[a] |
| Triangular | 6.0% (1.1% to 10.9%) | −2.1% (−7.2% to 3.0%) |
| Uniform | 7.3% (3.3% to 11.4%) | −1.7% (−6.0% to 2.7%) |

Abbreviations: CI = confidence interval; COPD = chronic obstructive pulmonary disease.

[a] Bias-corrected discontinuity estimate with data-driven bandwidth selection and robust standard errors.

**e-Table 10.** Polynomial Order and Effect Estimates

| Polynomial Order | | COPD Diagnosis | COPD Treatment |
|---|---|---|---|
| **Estimate** | **Bias** | **Effect (95% CI)**[a] | **Effect (95% CI)**[a] |
| 1 | 2 | 6.0% (1.1% to 10.9%) | −2.1% (−7.2% to 3.0%) |
| 1 | 3 | 3.6% (−2.2% to 9.5%) | −2.8% (−9.2% to 3.6%) |
| 1 | 4 | 0.7% (−6.8% to 8.1%) | −2.3% (−9.5% to 5.0%) |
| 2 | 3 | 4.4% (−1.4% to 10.1%) | −2.7% (−8.8% to 3.4%) |
| 2 | 4 | 0.1% (−7.1% to 7.4%) | −2.2% (−9.5% to 5.0%) |
| 3 | 4 | 0.8% (−6.3% to 7.8%) | −2.5% (−9.3% to 4.3%) |

Abbreviations: CI = confidence interval; COPD = chronic obstructive pulmonary disease.

[a] Bias-corrected discontinuity estimate with data-driven bandwidth selection and robust standard errors.

**e-Table 11.** Bandwidth Selection Methods and Effect Estimates

| | COPD Diagnosis | COPD Treatment |
|---|---|---|
| **Bandwidth Selection** | **Effect (95% CI)**[a] | **Effect (95% CI)**[a] |
| One MSE-optimal bandwidth selector | 6.0% (1.1% to 10.9%) | −2.1% (−7.2% to 3.0%) |
| Two MSE-optimal bandwidth selectors | 6.9% (2.5% to 11.3%) | −2.4% (−6.9% to 2.0%) |
| One MSE-optimal bandwidth selector for sum of estimates | 5.9% (1.0% to 10.9%) | −2.1% (−7.2% to 3.0%) |
| One CER-optimal bandwidth selector | 3.3% (−2.3% to 8.9%) | −2.4% (−8.3% to 3.5%) |
| Two CER-optimal bandwidth selectors | 5.4% (0.4% to 10.5%) | −2.4% (−7.5% to 2.8%) |
| One CER-optimal bandwidth selector for sum of estimates | 3.2% (−2.5% to 8.9%) | −2.4% (−8.3% to 3.5%) |

The local average treatment effect is equal to the absolute difference in the percentage of patients diagnosed or treated for COPD at the 0.7 cutoff, estimated using a regression discontinuity design. Robust bias-corrected confidence intervals and associated $P$ values are reported. Abbreviations: CER = coverage error rate; CI = confidence interval; MSE = mean square error.

[a] Bias-corrected discontinuity estimate with data-driven bandwidth selection and robust standard errors.

**e-Table 12.** Variance-Covariance Matrix Estimators and Effect Estimates

| | COPD Diagnosis | COPD Treatment |
|---|---|---|
| **Matrix Estimator** | **Effect (95% CI)**[a] | **Effect (95% CI)**[a] |
| 1 Nearest Neighbor | 6.0% (1.2% to 10.8%) | −2.1% (−7.2% to 3.0%) |
| 2 Nearest Neighbors | 6.0% (1.2% to 10.9%) | −2.1% (−7.2% to 3.0%) |
| 3 Nearest Neighbors | 6.0% (1.1% to 10.9%) | −2.1% (−7.2% to 3.0%) |
| 4 Nearest Neighbors | 6.0% (1.1% to 10.9%) | −2.1% (−7.1% to 3.0%) |
| 5 Nearest Neighbors | 6.0% (1.1% to 10.9%) | −2.1% (−7.2% to 3.0%) |
| Plug-In Residuals | 6.0% (1.0% to 11.1%) | −2.1% (−7.2% to 3.0%) |

Abbreviations: CI = confidence interval; COPD = chronic obstructive pulmonary disease.

[a] Bias-corrected discontinuity estimate with data-driven bandwidth selection and robust standard errors.

**e-Figure 1.** CONSORT Diagram

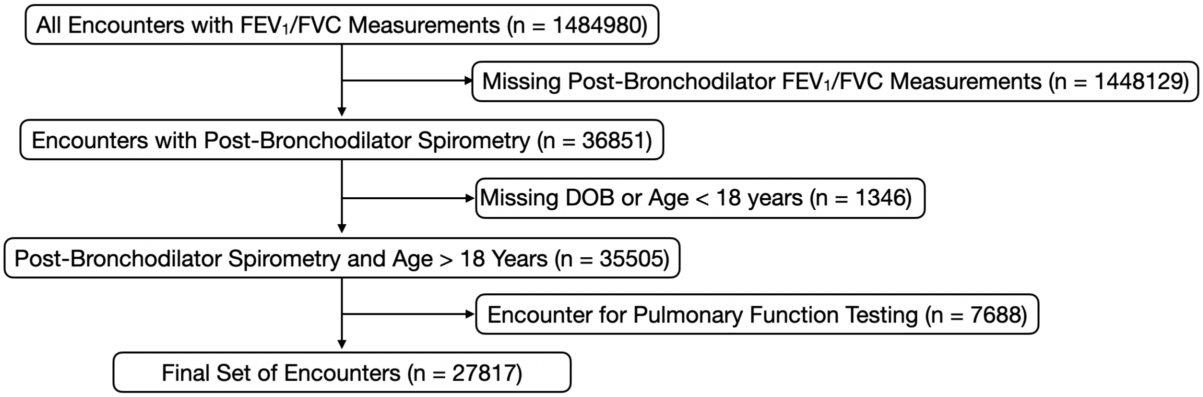

**e-Figure 2.** Distribution of Post-Bronchodilator $FEV_1/FVC$ Values

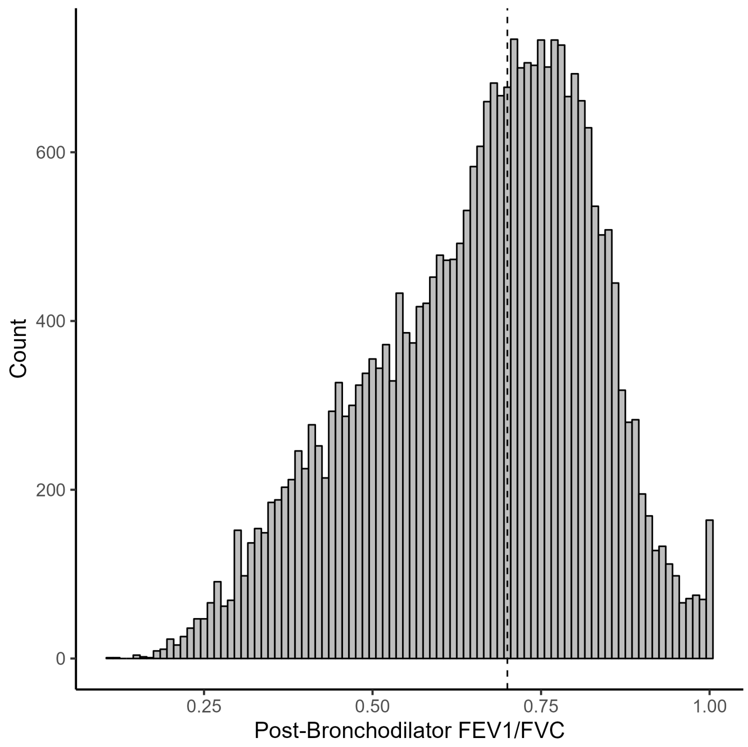

**e-Figure 3.** Distribution of Select Covariates at the Cutoff

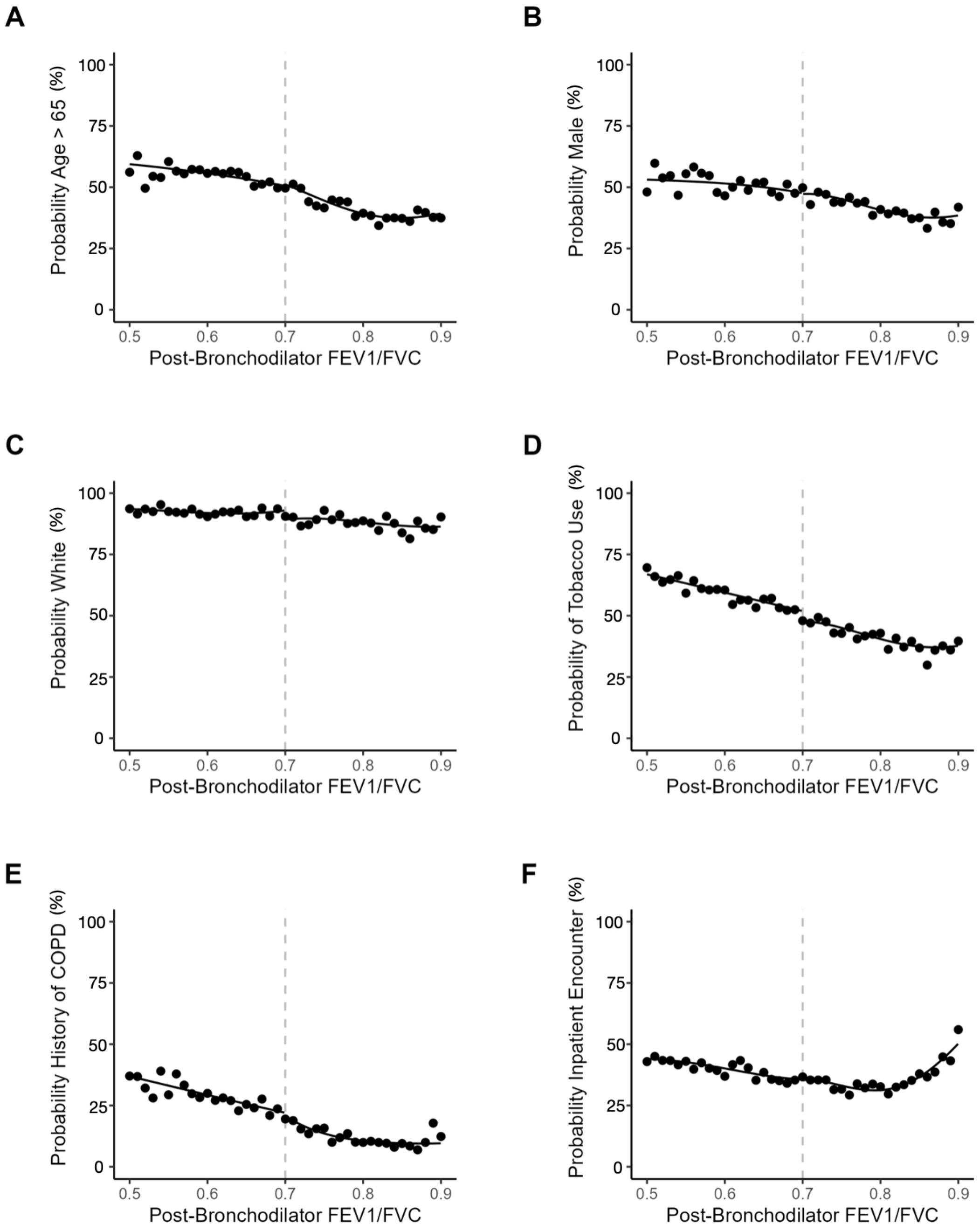

**e-Figure 4.** Association between Cutoff and Effect Estimates

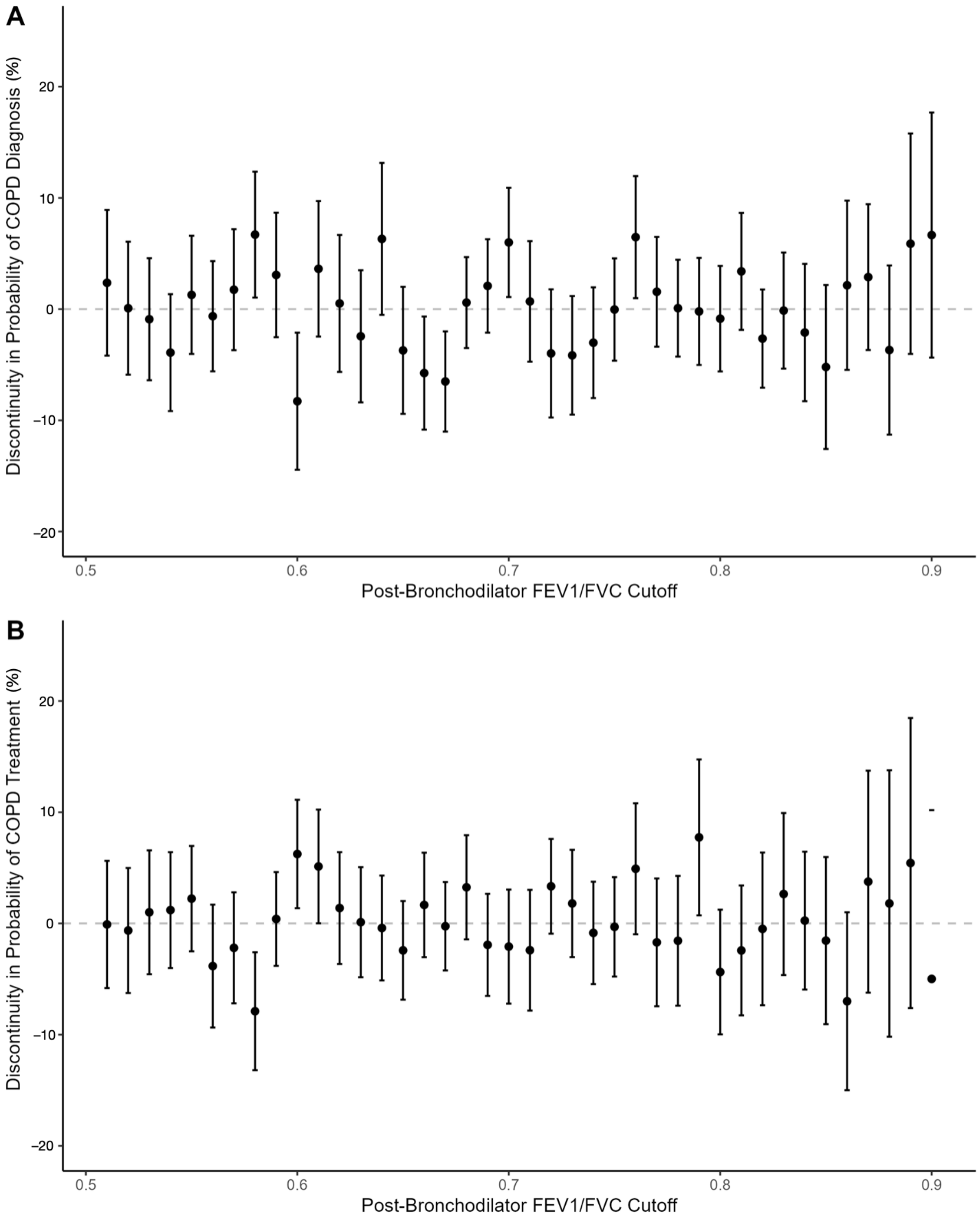

**e-Figure 5.** Association between Main Bandwidth and Effect Estimates

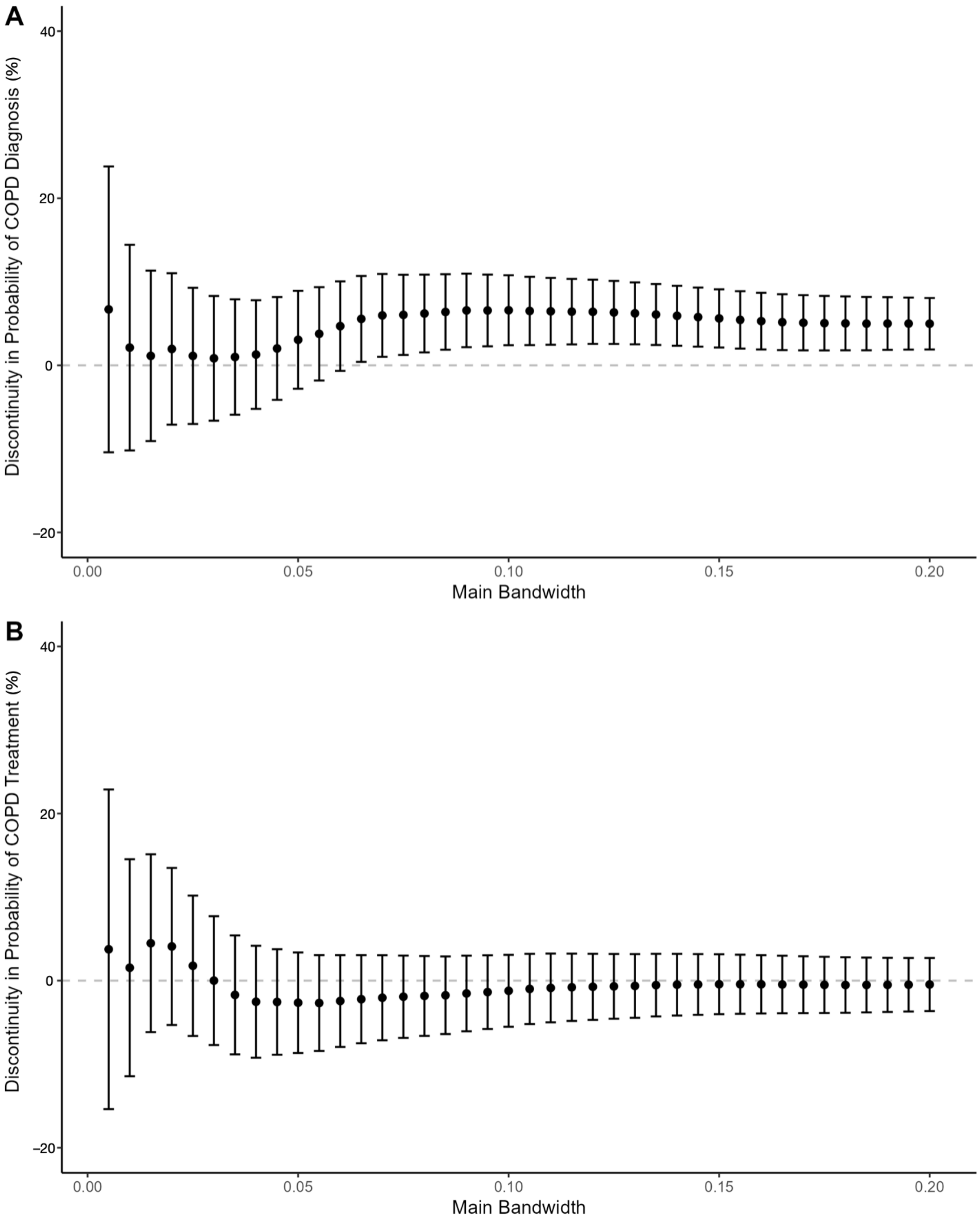

**e-Figure 6.** Association between Bias Bandwidth and Effect Estimates

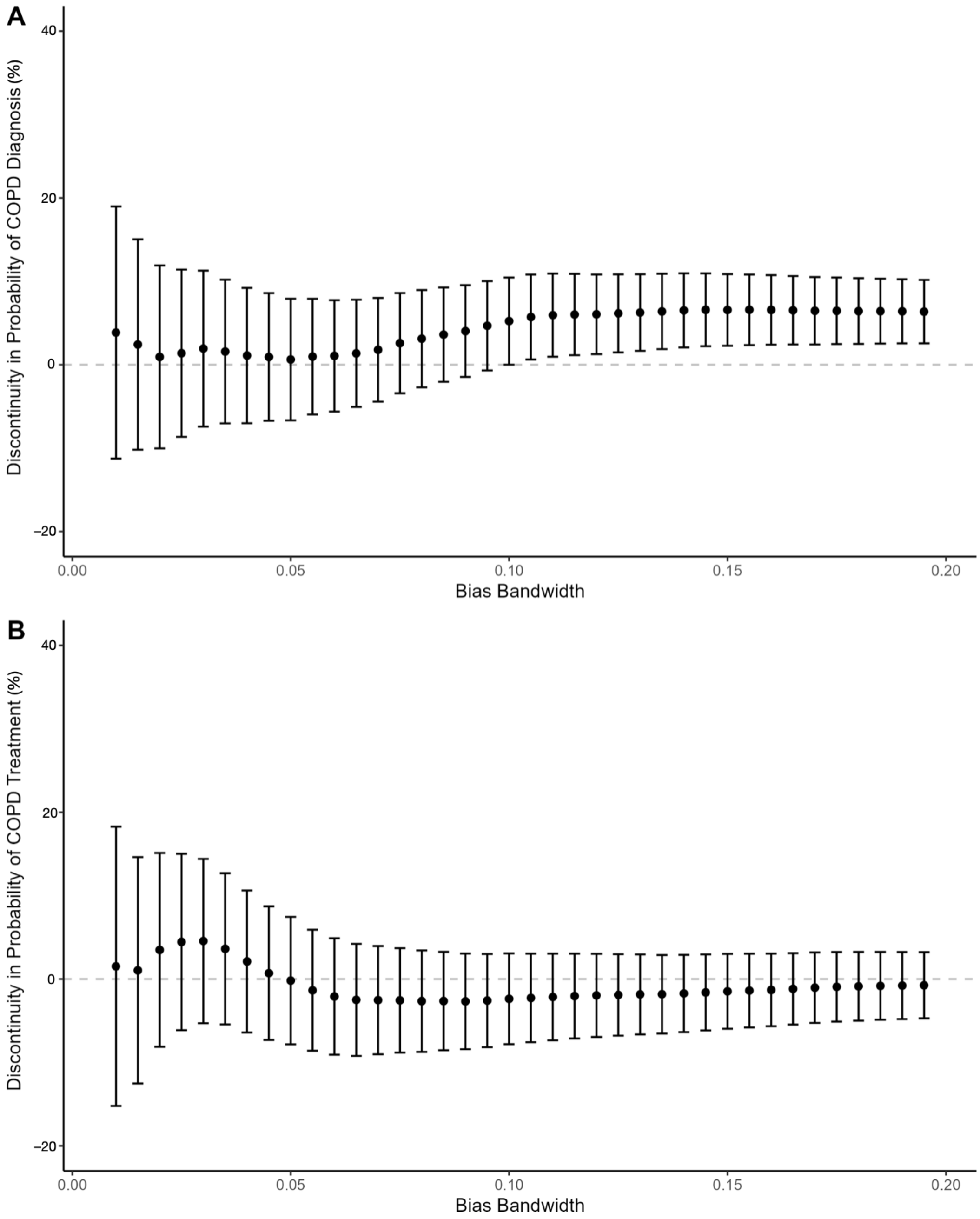

**e-Figure 7.** Sensitivity of Effect Estimate to Time to Diagnosis

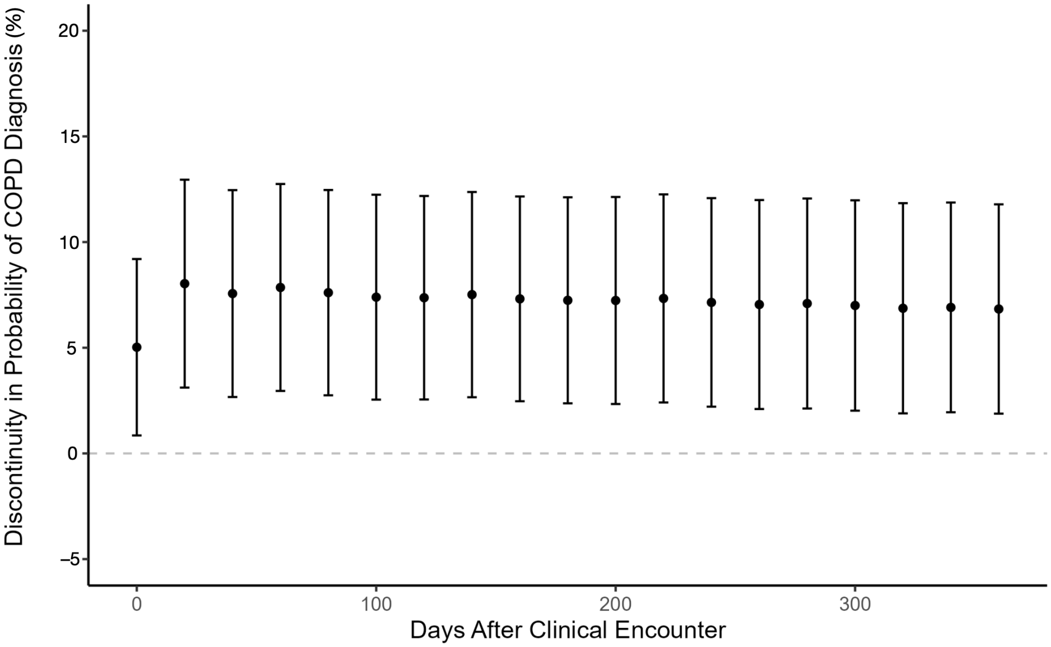

**e-Figure 8.** Sensitivity of Effect Estimate to Time to Treatment

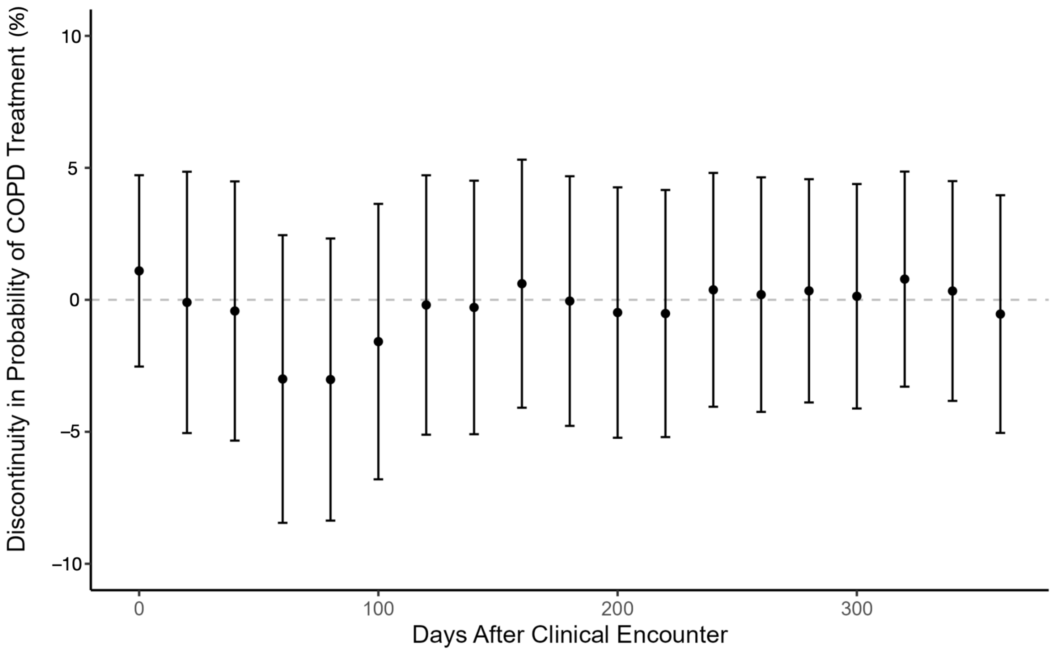
